# A Secondary Analysis Suggests That Repetitive Transcranial Magnetic Stimulation Applied to the Left Dorsolateral Prefrontal Cortex Reduces Cue-Induced-Craving in Treatment Seeking Participants with Cannabis Use Disorder

**DOI:** 10.1101/2025.01.16.25320690

**Authors:** Brendan L. Wong, Bohye Kim, Manpreet K. Singh, Jane P. Kim, Aimee L. McRae-Clark, Gregory L. Sahlem

## Abstract

**Background:** Studies across multiple addictions have suggested that repetitive transcranial magnetic stimulation (rTMS) applied to the left dorsolateral prefrontal cortex (L-DLPFC) reduces cue-induced-craving (CIC), however there are no studies in treatment seeking participants with cannabis use disorder (CUD). In this secondary analysis of a previously completed trial, we explore whether a multi-session course of rTMS reduces CIC in CUD.

**Methods:** Seventy-one participants with ≥moderate CUD (age=30.2±9.9;37.5% female) were randomized to twenty sessions of active or sham rTMS applied to the left DLPFC (20-sessions, Beam-F3; 10Hz) in a two-site, double-blind, sham-controlled, phase-2 trial where they also received motivational enhancement therapy. Participants rated their craving for cannabis via the short-form of the marijuana craving questionnaire (MCQ-SF) before and after a neutral and cannabis-cue presentation. Participants underwent assessment before (immediate-pre), after (immediate-post), and two-, and four-weeks following the course of rTMS.

**Results:** The MCQ-SF scores increased following the presentation of cannabis cues relative to neutral cues at the immediate-pre timepoint in both treatment groups (*p*<0.0001). Following study treatment, the percent increase in MCQ-SF following cues diverged between the active and sham groups with significantly reduced CIC in the active group at the two-week post time-point (5.8±7.1% sham group, 0.91±4.1% active group; *p*=0.02). Between-group effect sizes (Cohen’s d) were 0.24, 0.89, and 0.67 at the immediate-post, 2-week, and 4-week follow-up periods respectively.

**Conclusions:** L-DLPFC applied rTMS may reduce CIC in treatment seeking participants with CUD.

**Highlights:** Craving may be separable into tonic and phasic constructs

DLPFC applied rTMS did not effect tonic craving in a recent treatment trial for CUD

Data from that same trial suggests that DLPFC applied rTMS may have reduced phasic craving

## Introduction

Cue-reactivity refers to the physiologic response or increased desire to use a substance of choice (often referred to as craving) after seeing or being in the presence of that substance (or items/pictures/prompts to recall the use of that substance)^1–3^. Cue-reactivity can be measured behaviorally (cue-induced-craving), using physiologic recordings (such as heart-rate), or using neurophysiologic measurement (such as functional magnetic resonance imaging–fMRI). Clinically, a description of craving or cue-induced-craving is a harbinger of a return to use in individuals with substance use disorders, and this clinical observation is supported by an evidence base that prospectively links both spontaneous craving (also referred to as tonic-craving), and cue-induced-craving (also referred to as phasic-craving) toward future substance use^4–8^.

Pharmacotherapy studies have demonstrated that it is possible to engage both spontaneous and cue-induced-craving in human laboratory paradigms and clinical trials (showing that both of these aspects of craving can be engaged independently)^9–14^. Several studies in cannabis use disorder (CUD) have looked at various cue-reactivity paradigms^3^, though to date only one^12^ has looked at the pharmacologic engagement of cue-induced-craving in a human laboratory setting, and to our knowledge, no trial has attempted to engage cue-induced-craving in treatment-seeking participants in the context of a clinical trial. It is therefore unclear if any cue-induced-craving paradigm has clinical relevance, or whether it can be engaged by any intervention in CUD.

There is an expanding literature that reports on how applying repetitive Transcranial Magnetic Stimulation (rTMS) to various cortical targets effects craving^15^. The DLPFC in particular has been well studied using a variety of paradigms and findings suggest that in general DLPFC-applied-rTMS reduces spontaneous and cue-induced-craving, but there are negative reports of each as well. To date it is unclear if there is a differential effect in these craving constructs, though there may be somewhat more consistent effects of rTMS on cue-induced-craving. In CUD specifically, our group has found that a single-session of rTMS applied to the L-DLPFC decreased the purposefulness aspect of cannabis craving following the presentation of cues in a group of non-treatment-seeking participants and suggested decreased overall cue-induced craving^16^. In our larger multi-session clinical trial, we did not find a between group difference in spontaneous craving^17^ in treatment seeking participants.

We measured cannabis cue-induced-craving (CIC), as a secondary outcome in the above mentioned two-site, phase-2, double-blind, sham-controlled, randomized controlled trial testing the preliminary efficacy of rTMS applied to the L-DLPFC in CUD (primary outcomes have previously been reported^17^). We hypothesized that participants receiving active stimulation would have less cue-induced-craving (self-reported craving would go up less following the presentation of cannabis cues) than those participants receiving sham-rTMS. We further hypothesized that both baseline levels of cue-induced-craving, and the amount of reduction in cue-induced-craving following the course of rTMS would predict the number of weeks of abstinence and the number of days-per-week of cannabis use in the follow-up period, similar to previous findings^18^.

## Methods

This is an analysis of a secondary outcome collected as part of a larger phase-2 clinical trial. Detailed methods and primary results have already been published^17^. Briefly, the study was a sequential two-site, double-blind, randomized, sham-controlled clinical trial performed initially at the Medical University of South Carolina, and then at Stanford University. Participants were recruited through clinic referrals and social media advertisements. Participants underwent a structured screening and enrollment visit and were included if they met the following inclusion criteria: a) they were between the ages of 18 and 60 years; b) they met the Diagnostic and Statistical Manual for Mental Disorders—DSM-5–criteria for ≥moderate cannabis use disorder; c) they had a desire to quit or reduce cannabis use; and d) they had a positive urine drug test for cannabis. Participants were excluded if: a) they were pregnant or breast-feeding; b) they met DSM-5-criteria for another ≥moderate substance use disorder (other than nicotine use disorder); c) they were regularly taking medications with central nervous system effects; d) they had a history of psychotic disorder, bipolar disorder, or any other psychiatric condition requiring acute treatment; e) they had a Hamilton Rating Scale for Depression—HRSD_24_ score greater than 10 indicating clinically relevant depressive symptoms; f) they had a history of dementia or other cognitive impairment; g) they had active suicidal ideation or a suicide attempt within the past 90-days; h) they had any contraindications to receiving rTMS or MRI, or; i) they had any unstable general medical condition. After enrolling, participants underwent study-rTMS over approximately five-weeks, where they received active or sham rTMS (using MagVenture’s electronic sham-system) on ten treatment visits occurring approximately twice each week. At each visit participants received two rTMS sessions (10Hz, Beam-F3, 5-seconds on, 10-seconds off, 4000 pulses, 120% rMT, delivered in the presence of cannabis cues) with a 30-minute inter-session-interval (totaling 20 rTMS study-treatment sessions). Participants additionally received a three-session motivational enhancement therapy (MET) intervention^19^ after approximately their 1st, 3rd, and 5th treatment sessions. Participants then returned for a two-week and four-week follow-up visit, scheduled after the final study-treatment visit, for follow-up questionnaires and assessments.

We measured cue-induced-craving using a validated paradigm (see **Figure-1**), similar to one described previously^20^, immediately before participants’ first rTMS visit (immediate-pre), immediately before their final rTMS visit (immediate-post), and then two- and four-weeks after the course of rTMS was delivered. The short form of the marijuana craving questionnaire^21^ was administered verbally at baseline (prior to either neutral or cannabis cue presentations), following a neutral cue presentation, and then following a cannabis cue presentation. In the neutral presentation, participants listened to an audio script prompting them to remember a pleasant experience they had at the beach. Participants then handled physical neutral cues consisting of paperclips, a pencil, a dry-erase marker, yellow mini notepads, Lipton tea bags, and tanbark (for an olfactory cue) for three-minutes. In the cannabis cue presentation, participants listened to an audio script prompting them to think about a recent pleasant experience they had when using cannabis. Participants then handled cannabis cues consisting of rolling papers, a vape-pen, a pack of mock blunts, a blunt container, a fake joint, artificial-marijuana with essential-oil of cannabis as an olfactory cue, and a glass pipe, for three-minutes.

**Figure-1:**
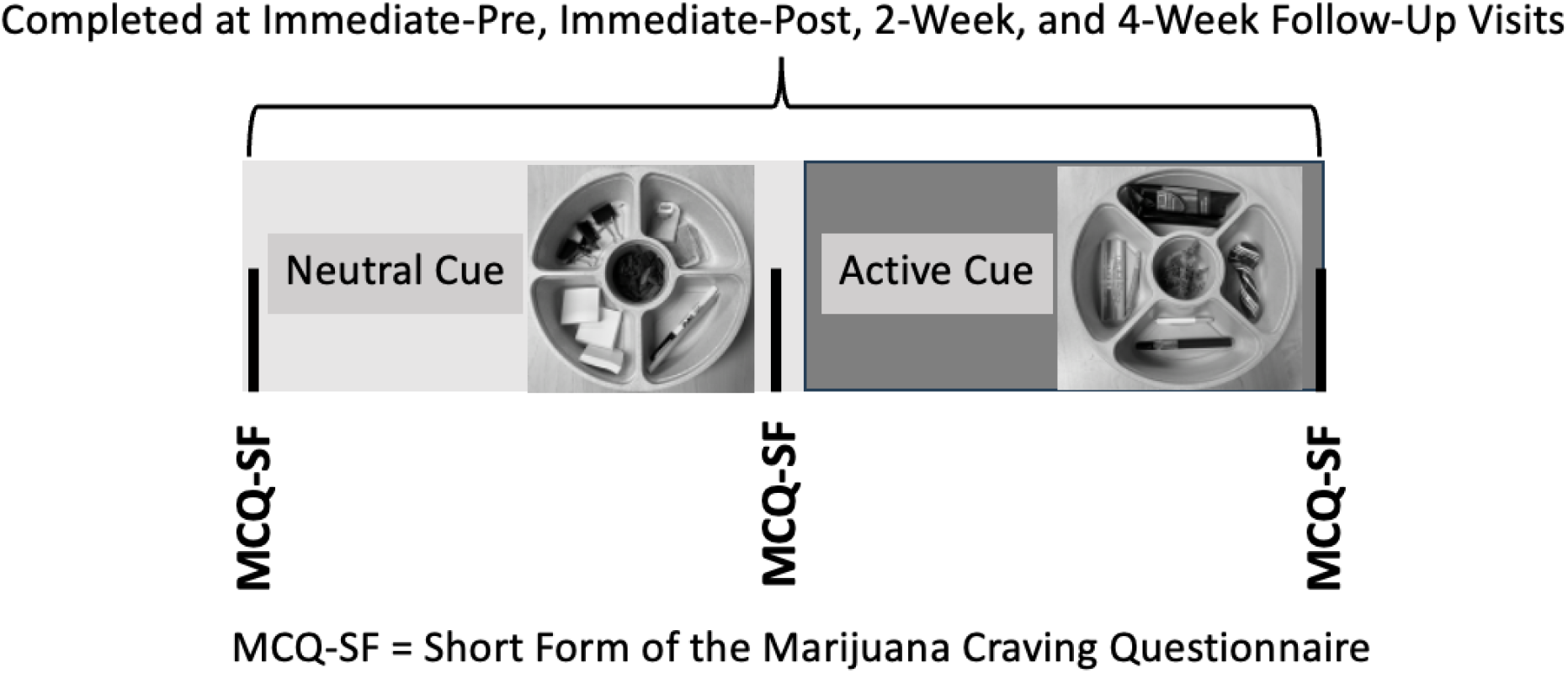
General flow diagram of cue paradigm.

The short-form of the marijuana craving questionnaire is a 12-item questionnaire where participants rate their agreement or disagreement on various items (1-’Strongly Disagree’, 4-’Neutral’, and 7-’Strongly Agree’). All items are summed for a total score (ranging from 12-84), and there are four subscales (each with scores ranging from 3-21) that further partition types of cannabis craving into compulsivity, emotionality, expectancy, and purposefulness.

### Analysis

Our primary hypothesis was that the group who received active rTMS would have a reduction in cue-induced-craving (the amount the MCQ-SF increased between post-neutral and post-cue assessments) relative to the group who received sham-stimulation. We both looked at the trajectory of cue-induced-craving (whether the active group had progressively less cue-induced-craving relative to the sham-group over time), as well as the level of cue-induced-craving at each time point (independent of the other time-points). To examine the between-group difference in the trajectory of cue-induced-craving, we defined our dependent variables as the MCQ-SF change score from the post-neutral to the post-cue assessment (post-cue score minus the post-neutral score; primary outcome) and the MCQ-SF change score from the baseline to the post-cue assessment (post-cue score minus the baseline score; secondary outcome). A linear mixed-effects model was fit, with the change scores serving as dependent variables and treatment condition, time, their interaction, and site serving as independent variables. To examine the between group difference in cue-induced-craving at each time-point, we performed a two-sample paired t-test for each of the four time-points using change scores as defined above. We also tested whether the change scores of cue-induced-craving (post-cue minus post-neutral, post-cue minus baseline) at the immediate-pre or the amount of change in the change scores from the immediate-pre to the immediate-post) were associated with the number of weeks of abstinence or days-per-week of cannabis use in the four-week follow-up period. For this, we fit linear models where dependent variables were the number of weeks of abstinence or days-per-week of cannabis use in the four-week follow-up period and independent variables were cue-induced-craving change scores (or the amount of change in the change scores from the immediate-pre to the immediate-post), treatment condition, and site. Each of the above analyses was repeated for the four MCQ-SF subscales (i.e. compulsivity, emotionality, expectancy, and purposefulness). Statistical significance was defined based on two-tailed tests with an alpha of 0.05 and reported without correcting for multiple comparisons in this preliminary investigation. We additionally reported effect sizes using Cohen’s *d*.

## Results

A total of 71 participants were included in this secondary analysis with n=37 receiving active stimulation and n=34 receiving sham stimulation. Baseline participant characteristics and demographics are presented in **Table-1**. A total of 52 participants (73%) completed study-treatment (28-active, 24-sham) and contributed to the immediate-post data-point. Forty-two participants (59%) contributed data to the 2- and 4-week follow-up assessments (23-active, 18-sham). We began measuring cue-induced-craving at these later time-points partially through the trial which is why the full sample of participants completing the trial did not contribute to the later time-points. In the combined sample (N=71), at the immediate-pre time-point, total MCQ-SF scores significantly decreased from baseline (45.5±17.3) to the post-neutral assessment (41.1±17.2), (T(67)= 4.74, p<0.0001). Scores then increased significantly following the presentation of cannabis cues (46.2±20.2), compared to the neutral assessment (T(68)= −4.57, p<0.0001). The MCQ-SF score did not differ significantly between the baseline and post-cue timepoint, (T(68)= −0.03, p=0.97). Each sub-scale followed a similar trend (see **Table-2** below).

**Table-1:** Baseline participant demographics and characteristics. All values are reported ± Standard Deviations. Cannabis use variables are reported for the 28-days prior to the screening and enrollment visit. There were no significant between-group differences.

|  | Full Sample | Active | Sham |
| --- | --- | --- | --- |
| Age (Years) | 30.2±9.9 | 29.8±9.9 | 30.7±10.0 |
| Sex | 62.5% Male<br>37.5% Female | 67.57% Male<br>32.4% Female | 57.14% Male<br>42.9% Female |
| Number of DSM-5 CUD categories met | 8.5±1.5 | 8.5±1.7 | 8.4±1.3 |
| CUDIT-R score | 21.4±5.1 | 21.3±4.5 | 21.6±5.7 |
| Marijuana Problem Scale (MPS) Score | 9.9±6.8 | 10.3±6.8 | 9.4±6.9 |
| Days of cannabis use (past 28-days) | 26.3±3.9 | 26.2±4.7 | 26.3±2.9 |
| Average number of cannabis use-sessions-per-day (past 28-days) | 4.4±3.3 | 4.2±2.9 | 4.6±3.7 |

DSM-5 = 5<sup>th</sup> edition of the Diagnostic and Statistical Manual of Mental Disorders; CUDIT-R = Revised edition of the Cannabis Use Disorder Identification Test.

**Table-2:** MCQ-SF scores at the immediate-pre for the full sample (participants assigned to active or sham.

|  | Baseline | Post-Neutral | Post-Cue | Significance Baseline to Neutral | Significance Neutral to Cue | Significance Baseline to Cue |
| --- | --- | --- | --- | --- | --- | --- |
| MCQ-SF total | 45.5±17.3 | 41.1±17.2 | 46.2±20.2 | <0.0001 | <0.0001 | 0.97 |
| Compulsivity | 8.8±4.9 | 7.6±4.4 | 9.2±5.5 | 0.0008 | 0.0001 | 0.44 |
| Emotionality | 10±5.4 | 8.9±5.3 | 10.1±6.1 | 0.008 | 0.001 | 0.91 |
| Expectancy | 12.4±5.2 | 11.4±5.3 | 12.2±5.7 | 0.0009 | 0.004 | 0.27 |
| Purposefulness | 14.5±5.1 | 13.1±5.5 | 14.7±5.7 | 0.0002 | 0.0003 | 0.67 |

There were no significant treatment-by-time effects for cue-induced-craving (MCQ-SF) either between post-neutral and post-cue presentations (□ = −0.23, p=0.41) or, baseline and post-cue presentations (□ = −0.49, p=0.12), though the active group showed numerically greater reductions than did the sham group. Similar trends held for the MCQ-SF subscales in both time trajectories (see **Table-3 and Supplemental-Table-1**).

**Table-3:** Treatment-by-time effects of cue-induced-craving, post-cue minus post-neutral scores.

|  | Significance (Treatment*Time) |
| --- | --- |
| Cue-Neutral |  |
| MCQ-SF total (Cue-Neutral) | $\square = -0.23, p=0.41$ |
| Compulsivity (Cue-Neutral) | $\square = -0.08, p=0.40$ |
| Emotionality (Cue-Neutral) | $\square = 0.03, p=0.73$ |
| Expectancy (Cue-Neutral) | $\square = -0.12, p=0.15$ |
| Purposefulness (Cue-Neutral) | $\square = -0.16, p=0.13$ |

Note: The □ indicates treatment by time effect.

**Table-4:** Percent change in MCQ-SF score between post-neutral and post-cue craving scores.

|  |  | Immediate-Pre | Immediate-Post | 2-week FU | 4-week FU |
| --- | --- | --- | --- | --- | --- |
| MCQ-SF Total | Active | 7.02±13.50% | 2.67±5.09% | 0.91±4.09% | 0.79±2.99% |
|  | Sham | 7.87±13.70% | 4.51±9.71% | 5.83±7.14% | 3.80±6.15% |
| Significance and effect size |  | p=0.80, d=-0.06 | p=0.41, d=0.24 | p=0.02, d=0.90 | p=0.09, d=0.67 |
| Compulsivity | Active | 8.80±16.7% | 1.71±9.53% | 1.69±4.57% | 0.00±2.37% |
|  | Sham | 8.99±19.6% | 4.17±11.0% | 4.17±9.83% | 4.51±6.48% |
| Significance and effect size |  | p=0.97, d=-0.01 | p=0.40, d=0.24 | p=0.36, d=0.34 | p=0.02, d=1.00 |
| Emotionality | Active | 5.09±16.9% | 2.47±4.95% | 0.24±5.16% | -0.24±4.88% |
|  | Sham | 9.26±18.4% | 2.78±7.86% | 4.17±9.83% | 2.78±11.5% |
| Significance and effect size |  | p=0.33, d=-0.24 | p=0.87, d=0.05 | p=0.16, d=0.53 | p=0.33, d=0.37 |
| Expectancy | Active | 4.48±15.9% | 1.23±5.41% | -0.24±7.39% | -0.24±7.58% |
|  | Sham | 4.74±8.98% | 4.86±12.0% | 3.82±9.01% | 5.93±15.9% |
| Significance and effect size |  | p=0.93, d=-0.02 | p=0.18, d=0.40 | p=0.13, d=0.50 | p=0.18, d=0.53 |
| Purposefulness | Active | 9.72±20.3% | 4.94±12.7% | 1.93±9.11% | 3.62±5.19% |
|  | Sham | 7.52±17.9% | 6.25±14.7% | 9.63±11.2% | 8.68±20.9% |
| Significance and effect size |  | p=0.63, d=0.11 | p=0.73, d=0.10 | p=0.03, d=0.77 | p=0.36, d=0.36 |

At the immediate-pre time-point, the active and sham groups had similar levels of cue-induced-craving. Following study-rTMS, there were general trends that the active-rTMS group had reduced cue-induced-craving relative to the sham-rTMS group, progressing from initially small toward larger effect sizes at subsequent follow-ups (between-group effect sizes were Cohen’s *d* = 0.24, 0.89, and 0.67 at the immediate-post, 2-week, and 4-week follow-up periods respectively). The numeric differences were present consistently in both cue minus neutral comparisons (primary), and cue minus baseline comparisons (secondary), and within each of the subscales (see **Figures-2a-e and Supplemental-Figures-1a-e**). There were significant group differences in the cue minus neutral total-score comparison at the 2-week post-treatment timepoint (T(20)=2.42, p=0.02, d=0.9), the cue minus neutral compulsivity comparison at the 4-week post-treatment timepoint (T(18)=2.66, p=0.02, d=1.0), the cue minus neutral purposefulness comparison at the 2-week post-treatment timepoint (T(36)=2.32, p=0.03, d=0.77), and the cue minus baseline compulsivity comparison at the 4-week post timepoint (T(37)=2.08, p=0.04, d=0.68), though each of these results were secondary analyses and not corrected for multiple comparisons in this preliminary study.

**Figures-2a-e.**
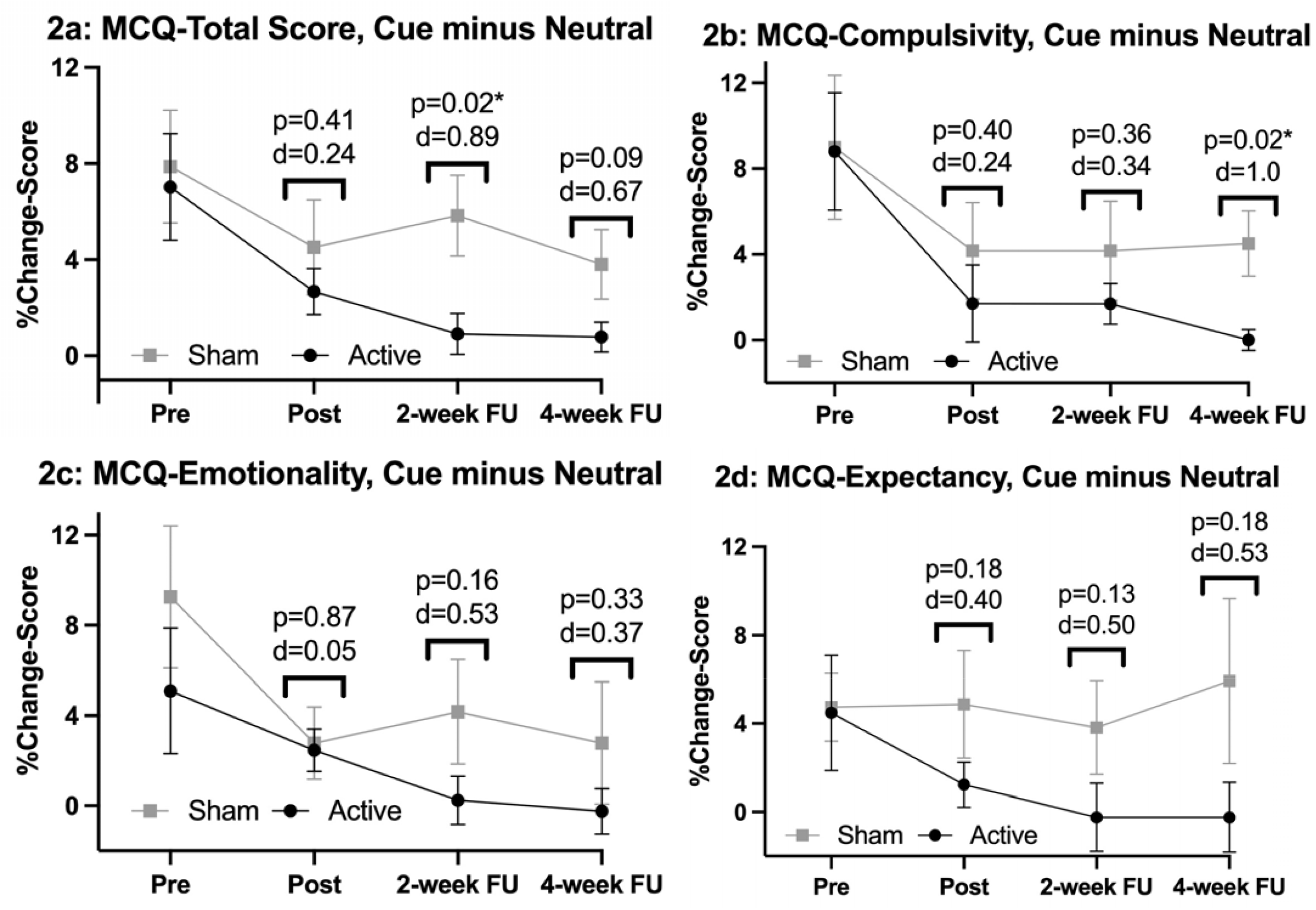
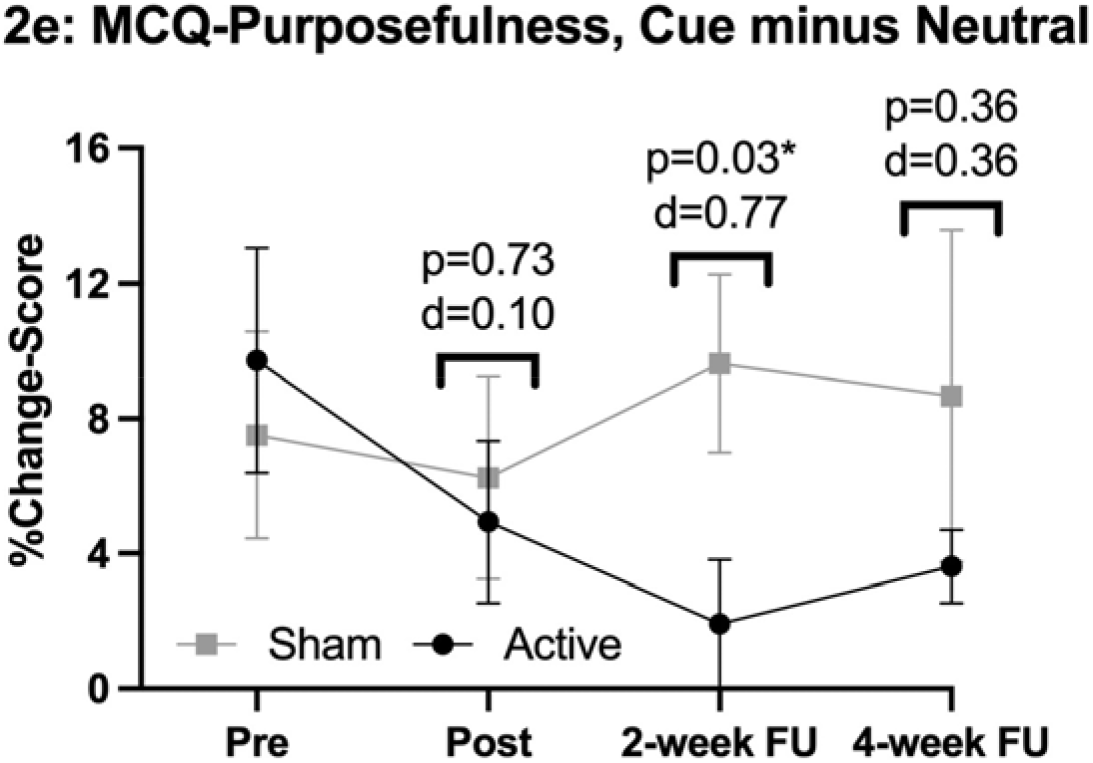
Cue-Induced Craving over time, Cue minus Neutral, in Total Score (2a), Compulsivity (2b), Emotionality (2c), Expectancy (2d), and Purposefulness (2e): These images depict the change in behavioral cue-induced-craving over time when subtracting the MCQ-SF score following the presentation of cannabis-cues from the MCQ-SF score following the presentation of neutral-cues. Error bars represent the Standard Error of the Mean (SEM), and *p*-values and Cohen’s d values are for each individual time point.

Neither the cue-induced-craving change scores (post-cue minus post-neutral, post-cue minus baseline) at the immediate-pre nor the amount of change in the cue-induced-craving change scores from the immediate-pre to the immediate-post time points were associated with either clinical outcome in the follow-up period (weeks of abstinence or days-per-week of cannabis use).

## Discussion

In this secondary analysis of a two-site, randomized-controlled clinical trial of prefrontally-applied rTMS for the treatment of cannabis use disorder we found that those participants who received active-rTMS had suggestions of reduced cue-induced-craving following a course of treatment relative to the participants receiving sham-rTMS, with medium effect sizes. Participants receiving active-rTMS became increasingly less cue-reactive over time; however, this effect failed to reach statistical significance between groups. Prospective study is needed with a larger sample size to determine whether this effect is present. We discuss our findings in relation to the published literature below and outline the strengths and limitations of this preliminary work.

Our preliminary findings suggest that rTMS applied to the L-DLPFC might be a promising approach to reduce cue-induced-craving in cannabis use disorder, and to our knowledge this is the first such intervention to hold promise in this symptom domain. In our earlier work^16^, we found that a single-session of rTMS may reduce cue-induced-craving. However, the effect was only observed in the purposefulness subscale, whereas in the current investigation, we observed numerical differences in all MCQ-SF subscales that persisted 4-weeks following study-rTMS, strengthening the possibility of a meaningful and durable effect. These data complement our finding that L-DLPFC applied rTMS did not have an effect on spontaneous craving^17^. It is unclear why we observed suggestions of between group effects of rTMS on cue-induced-craving but not spontaneous-craving. It is possible that the way we measured craving impacted the outcome, such that the MCQ-SF was better at measuring more immediate changes in craving then more longitudinal assessments of spontaneous-craving, or that spontaneous-craving is simply a different construct than cue-induced-craving. Indeed, in both human laboratory and clinical trials varenicline has differentially reduced spontaneous-craving while not effecting cue-induced-craving^9,11^. It is possible that DLPFC applied rTMS effects the opposite symptom domain, decreasing cue-induced-craving more robustly than spontaneous-craving, which would be supported by the regulatory role of the DLPFC on incentive-salience structures^22^. It is unclear why we did not replicate previous findings that suggested baseline cue-induced-craving predicts substance use, or previous findings that the level cue-induced-craving is reduced correlates with use outcomes. Small sample size may have played a role in our lack of concordant findings.

Though promising, our findings must be viewed in the context of some limitations. As previously noted, this was a phase-2 trial with a medium sample size. Second, our analysis was exploratory and did not correct for multiple comparisons. We also had incomplete retention with delayed introduction of the behavioral cue-reactivity paradigm at the two- and four-week follow-up visits, with 58% of the initial sample contributing data to the time-points where the effect sizes were largest. As such, reported data should be viewed as hypothesis-generating rather than hypothesis-testing.

Despite these limitations, this is the first trial to assess cue-induced-craving in a cohort of treatment seeking participants with CUD in the context of a clinical trial, and our findings are promising in a symptom domain that universally relates to substance use across substance use disorders. Having an intervention that can alter this phasic cue-induced craving is a novel and potentially important contribution to the treatment armamentarium. Thus, continued study in this novel area is worthy. Future investigations should include larger sample-sizes, deliver more sessions of rTMS in an optimized treatment paradigm, further attempt to disentangle treatment effects on spontaneous versus cue-induced craving, and measure the impact of decreased cue-induced-craving on other clinical outcomes. Finally, exploring alternative rTMS treatment targets may further elucidate whether the L-DLPFC is the optimal target for cue-induced craving.

## Data Availability

All data produced in the present study are available upon reasonable request to the authors

## Acknowledgements

The authors would like to acknowledge the National Institutes of Health for providing funding for this project via grants K23DA043628 (PI: Sahlem, NIH/NIDA), K12DA031794 (Co-PI’s McRae-Clark and Gray, NIH/NIDA), and K24DA038240 (PI: McRae-Clark, NIH/NIDA). We would also like to acknowledge the contributions of Mark George, Nathaniel Baker, Nolan Williams, Andrew Manett, Ian Kratter, Edward Short, Therese Killeen, Margaret Caruso, Lauren Campbell, Irakli Kaloani, Brian Sherman, Tiffany Ford, and Ahmad Musleh.

## Supplemental Data

**Supplemental Table 1:** Treatment by time effects of cue-induced-craving, Post-Cue - Baseline.

|  | Significance (Treatment*Time) |
| --- | --- |
| Cue-Baseline |  |
| MCQ-SF total (Cue-Baseline) | $\square = -0.49$, $T(124) = -1.580.49$, $p=0.12$ |
| Compulsivity (Cue-Baseline) | $\square = -0.13$, $T(132) = -1.320.13$, $p=0.19$ |
| Emotionality (Cue-Baseline) | $\square = -0.08$, $T(125) = -0.7608$, $p=0.454$ |
| Expectancy (Cue-Baseline) | $\square = -0.16$, $T(191) = -1.770.16$, $p=0.08$ |
| Purposefulness (Cue-Baseline) | $\square = -0.13$, $T(155) = -1.17 -0.13$, $p=0.25$ |

Note: The □ indicates treatment by time effect.

**Supplemental Table 2:** MCQ-SF change score between-Condition between baseline and post cue craving scores.

|  |  | Immediate-Pre | Immediate-Post | 2-week FU | 4-week FU |
| --- | --- | --- | --- | --- | --- |
| MCQ-SF Total | Active | 1.00±15.3% | 1.50±5.37% | -0.79±6.12% | -0.60±3.40% |
|  | Sham | -0.97±16.7% | 3.99±9.47% | 4.08±11.4% | 3.15±8.02% |
| Significance and effect size |  | p=0.61, d=0.12 | p=0.26, d=0.33 | p=0.13, d=0.56 | p=0.10, d=0.66 |
| Compulsivity | Active | 2.93±19.6% | 1.92±8.45% | 0.48 ±9.61% | -1.69±7.76% |
|  | Sham | 0.98±23.1% | 3.47±8.94% | 6.94±15.2% | 3.13±6.08% |
| Significance and effect size |  | p=0.70, d=0.09 | p=0.53, d=0.18 | p=0.15, d=0.53 | p=0.04, d=0.68 |
| Emotionality | Active | 1.23±21.1% | 1.92±7.02% | -2.66±12.4% | -1.21±5.79% |
|  | Sham | -1.96±26.4% | 0.93±10.8% | 0.69±11.8% | 4.17±14.3% |
| Significance and effect size |  | p=0.58, d=0.1361 | p=0.70, d=-0.11 | p=0.40, d=0.28 | p=0.17, d=0.53 |
| Expectancy | Active | -1.23±16.0% | 0.86±7.96% | -0.73±8.43% | -0.97±8.48% |
|  | Sham | -2.86±14.1% | 4.17±10.3% | 2.43±9.72% | 4.44±17.6% |
| Significance and effect size |  | p=0.66, d=0.11 | p=0.21, d=0.36 | p=0.29, d=0.35 | p=0.28, d=0.42 |
| Purposefulness | Active | 1.08±20.4% | 1.28±7.59% | -0.24±11.9% | 1.45±7.15% |
|  | Sham | 0.98±19.7% | 7.41±15.5% | 6.25±16.8% | 6.60±17.7% |
| Significance and effect size |  | p=0.98, d=0.005 | p=0.09, d=0.51 | p=0.17, d=0.46 | p=0.28, d=0.41 |

**Supplemental Figures-1a-e.**
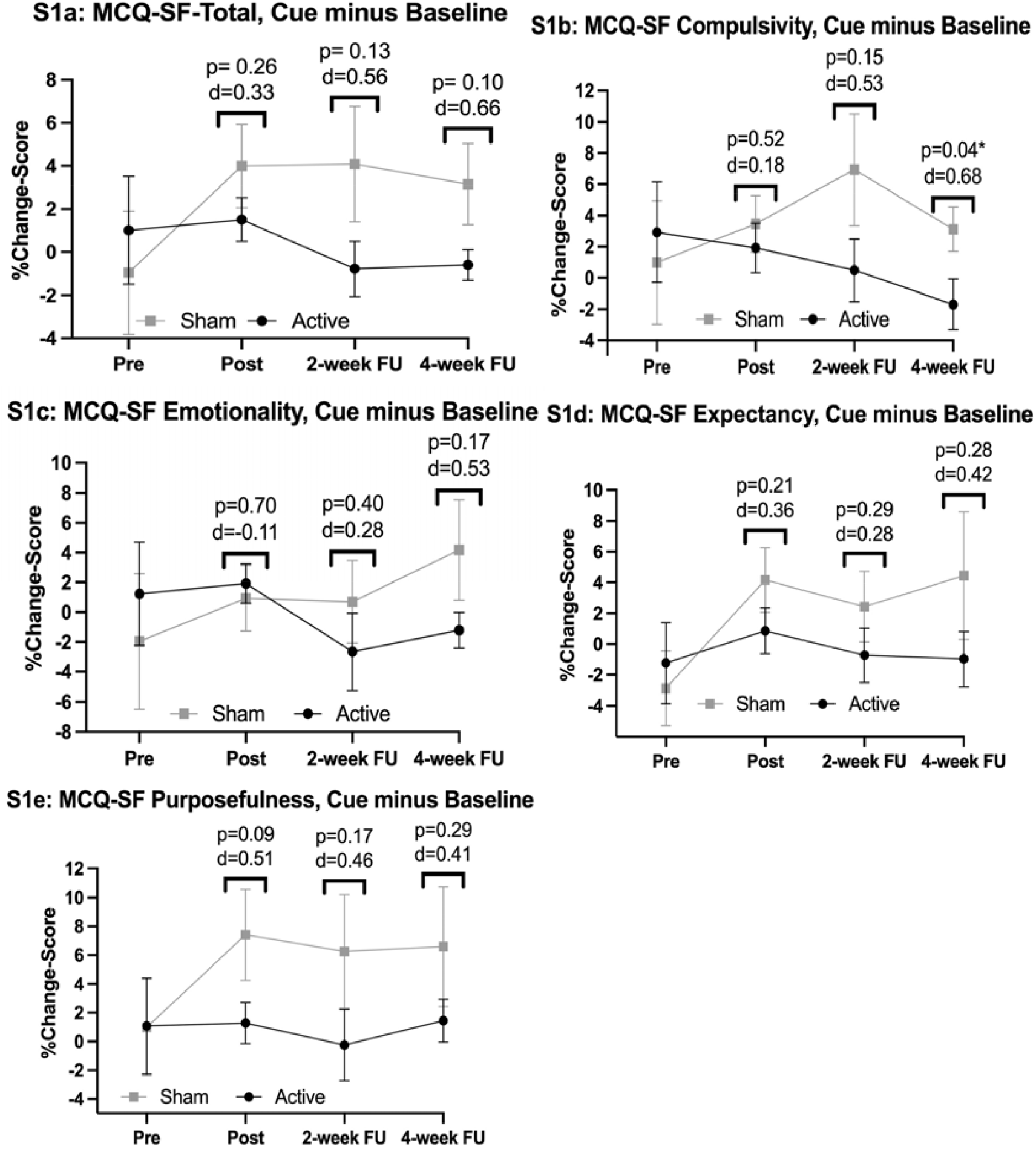
Cue-Induced-Craving over time, Cue minus Baseline in Total Score (S1a), Compulsivity (S1b), Emotionality (S1c), Expectancy (S1d), and Purposefulness (S1e): These images depict the change in behavioral cue-reactivity over time when subtracting the MCQ-SF score following the presentation of cannabis-cues from the baseline MCQ-SF score (prior to the presentation of any cues). Error bars represent the Standard Error of the Mean (SEM), and *p*-values and Cohen’s d values are for each individual time point.

## Notes

**Declaration of Interests:** MKS has received research support from the National Institutes of Health, the Patient Centered Outcomes Research Institute, and the Brain and Behavior Research Foundation. She is on a data safety monitoring board for a study funded by the National Institute of Mental Health. She has in the past 3 years consulted for AbbVie, Alkermes, Alto Neuroscience, Boehringer-Ingelheim, Johnson and Johnson, Karuna Therapeutics, Inc., and Neumora. She receives honoraria from the American Academy of Child and Adolescent Psychiatry and royalties from American Psychiatric Association Publishing. ALM has received research support from PleoPharma and has provided consultation to Indivior. GLS has collaborated with MagVenture and MECTA as part of investigator-initiated trials, consults for and has equity in the company Trial Catalyst, and has provided consultation to Indivior. None of the other authors have any relevant conflicts to disclose.

**Funding source:** This work was supported by the National Institutes of Health, Grant numbers: K23DA043628 (PI: Sahlem, NIH/NIDA), K12DA031794 (Co-PI’s McRae-Clark and Gray, NIH/NIDA), K24DA038240 (PI: McRae-Clark, NIH/NIDA).

### Competing Interest Statement

MKS has received research support from the National Institutes of Health, the Patient Centered Outcomes Research Institute, and the Brain and Behavior Research Foundation. She is on a data safety monitoring board for a study funded by the National Institute of Mental Health. She has in the past 3 years consulted for AbbVie, Alkermes, Alto Neuroscience, Boehringer-Ingelheim, Johnson and Johnson, Karuna Therapeutics, Inc., and Neumora. She receives honoraria from the American Academy of Child and Adolescent Psychiatry and royalties from American Psychiatric Association Publishing. ALM has received research support from PleoPharma and has provided consultation to Indivior. GLS has collaborated with MagVenture and MECTA as part of investigator-initiated trials, consults for and has equity in the company Trial Catalyst, and has provided consultation to Indivior. None of the other authors have any relevant conflicts to disclose.

### Clinical Trial

NCT03144232

### Funding Statement

This work was supported by the National Institutes of Health. Grant numbers: K23DA043628 (PI: Sahlem; NIH/NIDA); K12DA031794 (Co-PI's McRae-Clark and Gray; NIH/NIDA); K24DA038240 (PI: McRae-Clark; NIH/NIDA).

### Author Declarations

This work was approved by the Institutional Review Boards of both The Medical University of South Carolina and Stanford University. This is a secondary analysis paper and further details on informed consent and IRB approval are included in the referenced primary outcomes paper.

